# Addressing the ‘coin flip model’ and the role of ‘process of care’ variables in the analysis of TREWS

**DOI:** 10.1101/2022.09.13.22279688

**Authors:** Roy Adams, Katharine E Henry, Suchi Saria

## Abstract

Across two recent papers, Henry et al. (Nature Medicine, 2022) and Adams et al. (Nature Medicine, 2022) evaluated a deployed machine learning-based early warning system for sepsis, the Targeted Real-time Early Warning System (TREWS) for sepsis, finding that provider interactions with the tool were associated with reduced time to antibiotics and improved patient outcomes. In a subsequent commentary, Nemati et al. (medRxiv, 2022) assert that “the findings of Adams et al. are likely to be severely biased due to the failure to adjust for ‘processes of care’-related confounding factors.” In this response to Nemati et al., we argue that this conclusion is based on unrealistic assumptions about provider behavior that do not match the data reported in Adams et al. We further show that adjusting for ‘process of care’-related variables does not change the conclusions of Adams et al.

---

We first want to thank Nemati et al. for engaging with our work. We share their commitment to rigorous interrogation of the causal assumptions underlying observational clinical studies. In their response^1^ to our recent Nature Medicine article^2^, Nemati et al. assert that “the findings of Adams et al. are likely to be severely biased due to the failure to adjust for ‘processes of care’-related confounding factors.” Nemati et al. further claim that “the actual timing of the TREWS alert is of no consequence” in that we might find the same results using an alert that triggered for every patient at a random time. In this response, we first argue that these conclusions are based on strong and unrealistic assumptions about how providers respond to alerts. We then show that including ‘process of care’-related variables in our analysis – in addition to the demographic, environmental, and clinical variables already adjusted for in Adams et al. – does not substantively change the results of our primary analysis.

## The ‘coin flip model’

This main conclusions of Nemati et al. are motivated by the observation that, among sepsis patients, the presence of an early lactate measurement is strongly associated with improved patient outcomes. Thus, Nemati et al. reason, if prompt confirmation of a TREWS alert was associated with presence of an early lactate measurement, then the associations observed in Adams et al. may be the result of residual confounding. Nemati et al. verify this reasoning via a synthetic experiment in which they first generate an alert for every patient with a random delay relative to ED triage and then generate a hypothetical provider response that is strongly associated with a pre-alert lactate measurement above 1 mmol/L. They term this the ‘coin flip model’. Nemati et al. observe that, in this model, the synthetic provider response is associated with patient outcomes, but this association disappears when they adjust for the presence of a pre-alert lactate greater than 1 mmol/L. This type of reasoning has a long history in observational studies but is valid only to the extent that the hypothesized provider response matches reality. The results of Adams et al. are “likely to be severely biased,” as asserted by Nemati et al., only if providers are “likely” to behave as hypothesized by Nemati et al. No evidence to support this hypothesis is provided.

It is our belief that, if such an alert were deployed in practice, providers would be very unlikely to use it at all, much less to use it to reliably document suspected sepsis as hypothesized by Nemati et al. What Nemati et al. have *actually* observed is the well-documented result that early recognition of sepsis is associated with improved patient outcomes. Whereas, in previous works, “early” recognition was generally defined relative to ED triage^3,4^, Nemati et al. simply add an independent random offset to ED triage and make a similar observation. As we argue in Adams et al., ED triage (or a random timepoint thereafter) does not represent an actionable ‘time zero’ from which to measure time-to-recognition. One of the fundamental challenges of building a clinical alert system is balancing the earliness of the alerts against the precision of the alerts. That is, alerts that are too early may have low precision because there is not much information available on the patient. Alerts that are too late may have high precision (we have lots of information on the patient) but may not be actionable. The ‘coin flip model’ presents an alert that is neither early nor precise and hypothesizes that providers would respond to this alert in a meaningful way. Nemati et al. claim that the ‘coin flip model’ demonstrates that “the actual timing of the TREWS alert is of no consequence.” In practice, however, the timing and accuracy of the alert matters a great deal to providers and cannot be treated independently from the provider’s response to the alert.

## Adjusting for ‘process of care’ variables

The validity of the ‘coin flip model’ aside, Nemati et al. raise substantive concerns regarding two important sources of potential confounding:

***Environmental confounding*** may occur when environmental factors such as patient volume, alert volume, or time of day both delay response to the alert and adversely impact patient outcomes (e.g., by delaying care).

***‘Clinical suspicion’ confounding*** may occur when providers who already suspect sepsis at the time of the alert are both more likely to promptly confirm the alert and to promptly administer treatment, resulting in improved patient outcomes. Adjusting for this potential source of confounding requires adjusting for the providers level of suspicion at the time of the alert, which cannot be directly measured. Nemati et al. suggest the presence of a lactate measurement as an indicator that a provider may already suspect sepsis. Of note, the analysis published in Adams et al. already adjusts for a similar measure of abnormal lactate along with other laboratory measurements, vital signs, measures of severity, demographics, and comorbidities that may impact clinical suspicion or treatment of sepsis and may account for existing clinical suspicion. However, to check the sensitivity of our results, we adopt the suggestion from Nemati et al. Additionally, we include similar variables for both blood culture orders and fluids, early parts of a typical sepsis treatment bundle.

To test the sensitivity to these potential sources of confounding, we repeat our primary analyses. However, in addition to all the variables described in the “Adjustment variables” Section of Adams et al., we include:

### Environmental variables

1. Time of day discretized into 7am-3pm, 3pm-11pm, and 11pm-7am bins (corresponding to typical provider shifts)^5^.
2. Patient volume on the alert unit as measured by:
  a. Per hour admission rate over the last three hours minus the per hour admission rate over the last month. If fewer than two patients were admitted in the past three hours this feature was set to zero.
  b. Per hour admission rate over the last three hours divided by the per hour admission rate over the last month. If fewer than two patients were admitted in the past three hours this feature was set to zero.
  c. The CDF of a Poisson distribution with a mean of the per 3-hour admission rate over the last month evaluated at the number of admissions in the past three hours. This gives an approximate percentile of admission volume for the alert unit over the past three hours. If fewer than two patients were admitted in the past three hours this feature was set to zero.
3. Alert volume as measured by the number of alerts in the past three hours divided by the number of admissions in the past three hours. If the number of admissions in the past three hours is zero, this feature was set to zero.

### Clinical suspicion variables

4. For lactate, blood culture, and fluids orders:
  a. An indicator of whether an order occurred in the 24 hours prior to the alert.
  b. The time from the most recent order to the alert. This feature was set to zero if no order occurred in the preceding 24 hours.

## Results

Sample statistics for the newly included variables in the two exposure groups are shown in Table 1. As hypothesized by Nemati et al., we found higher rates of pre-alert lactate, blood culture, and fluids ordering in the study group than the comparison group. However, these associations are much smaller than those hypothesized in the ‘coin flip model’, providing further evidence that the model is not realistic. Additionally, the among patients who *had* received one of these orders in the 24 hours prior to the alert, the average time from order to alert was higher in the control arm. We also found statistically significant differences in the admission and alert volume variables, but, consistent with Henry et al., the magnitude of these differences was relatively small. Finally, study arm alerts were slightly less likely to have occurred between 11pm – 7am, but this difference was similarly small. Adjusted associations between prompt alert confirmation and patient outcomes are in Table 2. Though these associations are modestly smaller than those in Adams et al., the direction and statistical significance has not changed, and the 95% confidence intervals overlap substantially (∼82% overlap in the case of mortality).

**Table 1.** Sample statistics for newly included variables.

|  | Alert conf. w/in 3 hrs | Alert *not* conf. w/in 3 hrs | p-value |
| --- | --- | --- | --- |
| N | 4,220 | 2,657 | --- |
| Clinical suspicion |  |  |  |
| Lactate order before alert | 3,108 (73.6%) | 1,717 (64.6%) | < 0.001 |
| Hours since lactate order | 0.85 (1.2) | 1.14 (1.9) | < 0.001 |
| Culture order before alert | 2,958 (70.1%) | 1,366 (51.4%) | < 0.001 |
| Hours since culture order | 1.19 (2.7) | 1.94 (3.6) | < 0.001 |
| Fluids order before alert | 2,345 (55.6%) | 1,267 (47.7%) | < 0.001 |
| Hours since fluids order | 0.72 (1.2) | 1.45 (2.9) | < 0.001 |
| Environmental |  |  |  |
| 3hr adm. rate – 1mo adm. rate | 0.56 (0.4) | 0.49 (0.5) | < 0.001 |
| 3hr adm. rate / 1mo adm. rate | 0.55 (0.9) | 0.53 (0.9) | < 0.001 |
| 3hr adm. Poisson percentile | 1.32 (1.3) | 1.25 (1.5) | < 0.001 |
| Alerts per adm. (past 3 hrs) | 0.32 (0.4) | 0.30 (0.4) | < 0.001 |
| Time of day |  |  |  |
| 7am – 3pm | 814 (19.3%) | 496 (18.7%) | 0.544 |
| 3pm – 11pm | 2,046 (48.5%) | 1,243 (46.8%) | 0.177 |
| 11pm – 7am | 1,360 (32.2%) | 918 (34.6%) | 0.049 |

**Table 2.** Results with newly included variables.

|  | Adjusted risk difference or adjusted relative reduction | p-value § |
| --- | --- | --- |
| In-hospital mortality | ARD -2.73% [-4.52%, -1.06%] | 0.002 |
|  | ARR -15.18% [-23.73%, -6.26%] | 0.002 |
| SOFA progression at 72 hours ‡ | ARD -0.22 [-0.38, -0.07] | 0.005 |
| Median length of stay (hrs) † | ARD -9.10 [-15.60, -2.59] | 0.006 |
| Associations are reported as either an adjusted risk difference (ARD) or adjusted relative reduction (ARR) and presented as 'ARD/ARR [95% confidence interval]'.<br>‡ SOFA progression at 72 hours excludes patients who were discharged to hospice, left against medical advice, or were transferred to another acute care facility within 72 hours of the alert.<br>† Median length of stay was calculated only on patients who did not die in the hospital.<br>§ P-values for in-hospital mortality were based on non-parametric bootstrap resampling using 5,000 bootstrap samples. P-values for SOFA progression and median length of stay were based on t-tests. All tests were two-sided and were not adjusted for multiple comparisons. |  |  |

## Discussion

Our results show that, while Nemati et al. were correct to suspect an association between ‘clinical suspicion’ variables and provider response to the TREWS alert, this association was much smaller than they hypothesized. Adjusting for ‘process or care’ variables – in addition to the physiological and historical variables already adjusted for by Adams et al. – did not substantively impact the conclusions of Adams et al. The assertion that the results in Adams et al. were “likely to be severely biased” was premature and based on strong and unrealistic assumptions about how providers responded to TREWS alerts. While the importance of ‘clinical suspicion’ variables in predicting patient outcomes has been well documented^6,7^, the relationship between existing clinical suspicion and future treatment decisions is less well understood and likely merits further study.

## Data Availability

The data are not publicly available because they are from electronic health records approved for limited use by Johns Hopkins University investigators.

